# Association of axial length with retinal thickness and choroidal thickness in diabetic patients

**DOI:** 10.1101/2020.11.22.20236273

**Authors:** Sen Liu, Lanhua Wang, Wei Wang, Xia Gong, Yuting Li, Wangting Li, Xiaoling Liang, Yizhi Liu, Wenyong Huang

## Abstract

**Background:** Understanding the associations of axial length (AL) with retinal thickness (RT) and choroidal thickness (CT) at different subgrids among diabetic participants are of great important in exploring potential protective mechanism and pathogenesis of diabetic retinopathy (DR) in myopic eyes. Therefore, this study aimed to investigate the associations of AL with RT and CT among participants with type-2 diabetes mellitus (T2DM).

**Methods:** Participants with T2DM and registered with the government-monitored diabetes communities near Zhongshan Ophthalmic Center, Guangzhou, China, were consecutively invited to participate in the current study from October 2017 to April 2019. High-definition retina and choroid images of the macular area were obtained using swept-source optical coherence tomography.AL and other ocular biometrics were measured using Lenstar900. Linear regression models were used to assess relationships between AL and RT as well as CT.

**Results:** A total of 1378 participants with a mean age of 63.8±7.75 years and mean AL of 23.6±1.15 mm were included in the current study. In the multivariate linear regression models, AL was positively associated with the central RT (β=4.11 per mm increased in AL, 95%confidence interval (CI)=2.66 to 5.56, P<0.001), but negatively associated with the RT of the outer ring (β=-3.37 per mm increased in AL, 95%CI=-4.19 to -2.56, P<0.001).Longer AL tended to have thinker CTs in the central (β=-27.4 per mm increased in AL, 95%CI=-31.2 to -23.7, P<0.001),outer ring (β=-20.8 per mm increased in AL, 95%CI=-23.8 to -17.7, P<0.001) and inner ring (β=-24.6 per mm increased in AL, 95%CI=-28.1 to -21.1, P<0.001).

**Conclusions:** Myopic ocular elongation is accompanied by retinal thinning of the outer ring and retinal thickening of the foveal area. The CT of the macular area tended to become thinner with elongated AL among the diabetic subjects.

## Introduction

Diabetic retinopathy (DR) is a major microvascular complication of diabetes mellitus (DM) and can cause irreversible visual impairment and blindness. In fact, it is now the leading cause of blindness among working-age adults worldwide.^1, 2^ DR is becoming a great burden on healthcare systems and society. It is estimated that the number of people with DR will increase rapidly from 126.6 million in 2010 to 191 million by 2030 worldwide without prompt treatment.^2^ Despite tremendous progress in basic and clinical research, many questions regarding the risk factors and mechanisms of DR remain unanswered.

There is mounting evidence that longer axial length (AL) is a protective factor against DR.^3-6^ A possible explanation of this protective effect is that AL elongation is followed by chorioretinal degeneration, causing decreased blood flow and reduced metabolic demand, which in turn decrease the pathologic effect of hyperglycemia-induced microvasculature changes.^7, 8^ Thus, research on AL-related changes in the choroid and retina of diabetic patients would improve our understanding of the potential protective mechanism and pathogenesis of DR in myopic eyes.

With the introduction of spectral domain posterior segment optical coherence tomography (SD-OCT), it is now possible to evaluate the thickness characteristics of the retina and choroid noninvasively in living eyes. Numerous studies have investigated retinal thickness (RT) and choroidal thickness (CT) in diabetic eyes with or without DR.^9-12^ However, the existing studies on AL and choriodretinal thickness are limited and report conflicting results, as some studies indicate a negative association between AL and CT, whereas others report none. Moreover, studies on AL and choroid–retinal thickness in diabetic eyes are scarce.

The relatively low wavelength of SD-OCT hinders penetration through the retinal pigment epithelium (RPE), leading to the detection of a blurred choroid–scleral interface.^13^ Manual measurement at only one or a limited number of points^14^ may partly explain the aforementioned conflicting findings. Novel swept-source optical coherence tomography (SS-OCT) has a wavelength of 1,050 nm, which can provide a clear boundary of the choroid–scleral interface due to improved penetration of the RPE. Additionally, the automatic measurements, higher scanning speed (100,000 A scans/s) and resolution, as well as the three-dimensional (3D) models of the images make it more reliable than SD-OCT.^15^ To date, no studies have used SS-OCT to investigate the associations between AL, CT, and RT among diabetic patients. Therefore, using SS-OCT, this cross-sectional study was conducted to assess the associations of AL with RT and CT in different sub-regions among diabetic patients in China.

## Methods

### Study participants

This observational cross-sectional study was carried out in Zhongshan Ophthalmic Center (ZOC), Guangzhou, Southern China, from October 2017 to April 2019. Patients with type-2 diabetes mellitus (T2DM), aged between 30 and 80 years, and registered with the government-monitored diabetes communities near the ZOC were consecutively invited to participate in this study. The following exclusion criteria were applied: (1) history of severe systemic disease, (2) with a cognitive disorder or mental illness, (3) with van Herick value ≤ 25°, intraocular pressure (IOP) > 21 mm Hg, or previous history of primary angle-closure glaucoma that cannot be dilated, 4) history of cataract and other intraocular surgeries, (5) history of retinal laser photocoagulation, (6) epiretinal membrane, macular holes, age-related macular disease, retinal detachment, or other retinal disease, (7) best-corrected visual acuity (BCVA) < 20/200, and (8)missing data of ocular biometry, (9) poor OCT image quality (< 50, with eye movement, artefacts, and segmentation failure).

The study adhered to the tenets of the Declaration of Helsinki, and approval was obtained from the Institutional Review Board of the ZOC. Written informed consent was obtained from all participants.

### Ocular biometry other ocular examinations

Anterior chamber depth (ACD), lens thickness (LT), AL, central cornea thickness (CCT), cornea power (CP) were measured using Lenstar (LS900, Haag-Streit, Koeniz, Switzerland). The mean values of three measurements were recorded and used for the analysis. Uncorrected visual acuity (UCVA) and BCVA were measured with the Early Treatment Diabetic Retinopathy Study (ETDRS) visual chart at a distance of 4 m. Non-cycloplegic refractions were tested by an autorefractor (KR-8800, Topcon, Tokyo, Japan). IOP was measured by a non-contact tonometer (CT-1, Topcon, Tokyo, Japan) before mydriasis. Standardized 7-field fundus photos of both eyes were taken using a digital fundus camera (Canon CR-2, Tokyo, Japan) after mydriasis with 1% topicamide phenylephrine drops. Presence of DR was graded as R0, R1, R2, or R3 independently by two trained graders according to the United Kingdom National Diabetic Eye Screening Program (UK NDESP) guidelines.^16^

### Swept-source OCT measurements

Each participant underwent bilateral SS-OCT scanning (DRI-OCT-2 Triton, Topcon, Tokyo, Japan) by the same nurse to obtain high-definition images of the retina and choroid on the macular area after mydriasis. This SS-OCT device is a wavelength tunable laser (center wavelength: 1050 nm; tuning range: 100 nm) with a scan speed of 100,000 A scans/s, and it can yield an 8-μm axial resolution in tissue. The 6 × 6 mm raster scan model of the macular area was used to obtain 3D images. Then, an automated built-in layer segmentation software (Version 9.12.003.04) was used to analyze the images and automatically calculate the RTs and CTs in different subfields. The macular area was divided into nine subfields (the central, inner superior, inner nasal, inner inferior, inner temporal, outer superior, outer nasal, outer inferior, and outer temporal fields) and two rings (the inner ring at 1–3 mm and the outer ring at 3–6 mm) according to the ETDRS system with diameters of 1, 3, and 6 mm. The average values of the RTs and CTs in all the macular subfields were also analyzed.

### Systemic and laboratory examinations

Systolic (SBP) and diastolic (DBP) blood pressures, height, and weight were measured by the same trained nurse 15 min after sitting and shoes off. Body mass index (BMI) was calculated as the weight in kilograms divided by the square of the height in centimeters. Information on diabetes onset age and duration, current medication usage, history of systemic and ocular disease, and smoking and drinking statuses were collected by the same trained nurse to ensure reliability. A venous blood sample was used to test the concentration of hemoglobin A1C (HbA1c), total cholesterol (TC), triglycerides (TG), low density lipoprotein-cholesterol (LDL-C), high density lipoprotein-cholesterol (HDL-C), serum uric acid (UA), serum creatinine clearance rate (Scr), and C-reactive protein (CPR). The estimated glomerular filtration rate (GFR) (in mL/min/1.73 m^2^) was calculated according to the Chronic Kidney Disease Epidemiology Collaboration equation.^17^ The microalbuminuria level (in mg/dL) was assessed using a 50 mL midstream urine sample.

### Statistical analysis

Data of the right eye were analyzed in the current study. All statistical analyses were performed with Stata (ver. 12.0, Stata Corp, College Station, TX). Continuous variables were compared using Student’s *t* test or analysis of variance, while the Chi-squared test was employed for the comparison of proportions. The participants were further divided into four groups according to the AL quartiles. Univariable and multivariable line regression models were used to analyze potential associations of AL with RT and CT in different sub grids after adjusting for confounders. A *P* value of < 0.05 was considered to be statistically significant.

## Results

### Participants’ demographics and clinical features

A total of 1378(68.5%) participants were eligible for final analysis. Compared to those without DR, participants with DR tended to be younger (62.9 ± 8.40 vs. 64.0 ± 7.60, *P* = 0.046), male (51.3% vs. 41.4%, *P* = 0.002), had longer duration of diabetes (11.6 ± 7.34 vs. 8.16 ± 6.65, *P* < 0.001), and were more likely to use insulin (41.7% vs. 23.5%, *P* < 0.001). They also presented higher HbA1c (7.80 ± 1.79 vs. 6.80 ± 1.32, *P* < 0.001), higher SBP (136.6 ± 21.9 vs. 133.6 ± 19.7, *P* =0.033), higher level of microalbuminuria (11.1 ± 22.4 vs. 4.07 ± 14.9, *P* < 0.001), higher RT (239.1 ± 46.8 vs. 226.8 ± 27.3, *P* < 0.001), shorter AL (23.4 ± 1.13 vs. 23.6 ± 1.15, *P* < 0.001), shallower ACD (2.37 ± 0.34 vs. 2.43 ± 0.35, *P* = 0.009), and lower GFR (82.8 ± 17.9 vs. 87.0 ± 15.8, *P* < 0.001) (Table1).

**Table1.** Comparison of basic characteristic of participants with and without diabetic retinopathy

| Characteristic | No DR | Any DR | P-value |
| --- | --- | --- | --- |
| Total number,n (%) | 1148(83.3) | 230(16.7) |  |
| Age(years) | 64.0±7.60 | 62.9±8.40 | 0.046 |
| Female,n (%) | 684(59.6) | 112(48.7) | 0.002 |
| Duration of diabetes(years) | 8.16±6.65 | 11.6±7.34 | <0.001 |
| Use of insulin,n (%) | 268(23.5) | 96(41.7) | <0.001 |
| History of hypertension,n (%) | 638(55.6) | 126(54.8) | 0.825 |
| Smoker,n (%) | 191(24.5) | 42(30.7) | 0.124 |
| Drinker,n (%) | 191(24.5) | 42(29.2) | 0.238 |
| HbA1c (%) | 6.80±1.32 | 7.80±1.79 | <0.001 |
| BMI (kg/m<sup>2</sup>) | 24.7±3.27 | 24.5±3.35 | 0.437 |
| SBP(mmHg) | 133.6±19.7 | 136.6±21.9 | 0.033 |
| DBP(mmHg) | 71.0±11.9 | 70.7±11.4 | 0.660 |
| Cholesterol(mg/dL) | 187.3±39.5 | 185.2±41.5 | 0.480 |
| LDL-C(mg/dL) | 119.4±35.7 | 119.5±34.5 | 0.987 |
| HDL-C(mg/dL) | 51.4±15.7 | 50.0±15.5 | 0.219 |
| Triglycerides (mg/dL) | 206.8±148.6 | 210.0±152.5 | 0.769 |
| GFR(ml/min/1.73m<sup>2</sup>) | 87.0±15.8 | 82.8±17.9 | <0.001 |
| Microalbuminuria(mg/dL) | 4.07±14.9 | 11.1±22.4 | <0.001 |
| CPR(mg/dL) | 2.62±7.20 | 2.20±3.37 | 0.397 |
| CCT(μm) | 546.9±31.0 | 550.0±33.3 | 0.175 |
| CP(D) | 44.2±1.44 | 44.2±1.49 | 0.869 |
| AL(mm) | 23.6±1.15 | 23.4±1.13 | 0.027 |
| ACD(mm) | 2.43±0.35 | 2.37±0.34 | 0.009 |
| LT(mm) | 4.71±0.34 | 4.74±0.37 | 0.154 |
| SE(D) | 0.01±2.25 | 0.14±2.29 | 0.455 |
| Central retinal thickness(μm) | 226.8±27.3 | 239.1±46.8 | <0.001 |
| Central choroidal thickness(μm) | 206.1±85.0 | 209.1±80.7 | 0.635 |

All data presented as mean±standard deviation unless otherwise indicated
Abbreviations: DR = diabetic retinopathy; HbA1c = hemoglobin A1c; BMI = body mass
index; SBP =Systolic blood pressure; DBP =diastolic blood pressure; LDL-C=low density
lipoprotein; HDL-C=high density lipoprotein;GFR = glomerular filtration rate;
CPR=C-reactive protein; CCT=central cornea thickness;CP=cornea power;
D=dioptr;AL=axial length;ACD= anterior chamber depth;LT=lens thickness;SE=spherical
equivalent

The participants were further divided into four groups according to the AL quartiles. In general, participants with elongated AL tended to be male (*P* < 0.001) and exhibited higher BMI (*P* = 0.002), larger DBP (*P* = 0.002), higher UA (*P* < 0.001), lower cholesterol (*P* = 0.024), lower HDL-C (*P* < 0.001), deeper ACD (*P* < 0.001), thicker CCT(*P* < 0.001), smaller cornea power (*P* < 0.001) and LT(*P* < 0.001), less hyperopia (*P* < 0.001), and were less likely to have any DR (*P* = 0.013) (Table 2).

**Table2.** Comparison of basic characteristic stratified by AL quartile among diabetic participants

| Characteristic | Axial length(mm) |  |  |  | P value |
| --- | --- | --- | --- | --- | --- |
|  | First quartile<br>(<22.84) | Second quartile<br>(22.84~23.43) | Third quartile<br>(23.43~24.06) | Fourth quartile<br>(≥24.06) |  |
| Age (years) | 63.2±7.15 | 64.1±7.64 | 64.7±7.68 | 63.4±8.39 | 0.038 |
| Female,n (%) | 269(79.8) | 206(59.4) | 171(48.6) | 150(43.9) | <0.001 |
| Duration of diabetes(years) | 8.92±6.62 | 9.10±6.91 | 8.10±6.61 | 8.84±7.37 | 0.237 |
| HbA1c (%) | 7.07±1.53 | 6.91±1.40 | 6.96±1.52 | 6.94±1.36 | 0.521 |
| Use of insulin,n (%) | 97(28.8) | 89(25.8) | 81(23.1) | 97(28.6) | 0.257 |
| Smoker,n (%) | 56(25.9) | 71(29.5) | 59(25.1) | 47(20.8) | 0.198 |
| Drinker,n (%) | 53(24.5) | 70(29.1) | 57(24.3) | 51(22.6) | 0.410 |
| History of hypertension,n (%) | 182(54.0) | 180(51.9) | 203(57.7) | 199(58.2) | 0.281 |
| BMI (kg/m<sup>2</sup>) | 24.1±3.42 | 24.5±3.42 | 24.9±3.02 | 24.9±3.19 | 0.002 |
| SBP (mmHg) | 133.9±20.3 | 134.3±19.0 | 133.4±21.2 | 134.8±19.9 | 0.838 |
| DBP (mmHg) | 70.5±10.8 | 70.1±10.8 | 70.3±10.2 | 73.2±11.4 | 0.002 |
| Cholesterol(mg/dL) | 192.4±40.2 | 186.7±39.0 | 183.7±38.8 | 185.1±40.9 | 0.024 |
| LDL-C(mg/dL) | 123.5±36.2 | 117.9±34.5 | 118.2±34.7 | 118.4±36.5 | 0.118 |
| HDL-C(mmol/L) | 55.1±17.3 | 51.5±16.1 | 48.4±13.8 | 49.7±14.6 | <0.001 |
| Triglycerides (mg/dL) | 192.3±123.6 | 213.6±160.8 | 216.0±163.7 | 206.8±143.8 | 0.156 |
| GFR(ml/min/1.73m<sup>2</sup>) | 88.1±14.8 | 86.1±17.1 | 85.8±16.1 | 85.3±16.6 | 0.130 |
| Microalbuminuria(mg/dL) | 4.80±13.9 | 5.06±18.8 | 4.96±16.3 | 6.00±16.9 | 0.792 |
| CPR(mg/dL) | 2.03±2.11 | 2.65±6.70 | 2.60±4.04 | 2.91±10.8 | 0.375 |
| CCT(μm) | 542.0±29.9 | 546.9±32.5 | 547.1±30.0 | 553.6±32.1 | <0.001 |
| CP(D) | 45.4±1.20 | 44.4±1.09 | 43.7±1.09 | 43.2±1.35 | <0.001 |
| ACD (mm) | 2.21±0.32 | 2.36±0.28 | 2.49±0.31 | 2.62±0.35 | <0.001 |
| LT(mm) | 4.79±0.32 | 4.73±0.32 | 4.69±0.34 | 4.64±0.37 | <0.001 |
| SE (D) |  |  |  |  | <0.001 |
| Hyperopia | 232(69.7) | 200(58.5) | 168(48.3) | 68(20.2) |  |
| Emmetropia | 77(23.1) | 100(29.2) | 121(34.8) | 68(20.2) |  |
| Mild myopia | 24(7.21) | 36(10.5) | 51(14.7) | 88(26.1) |  |
| Moderate myopia | 0 | 6(1.75) | 8(2.30) | 73(21.7) |  |
| High myopia | 0 | 0 | 0 | 40(11.9) |  |
| Any DR | 64(19.0) | 72(20.8) | 51(14.5) | 43(12.6) | 0.013 |

All data presented as mean±standard deviation unless otherwise indicated.
Abbreviations: HbA1c = hemoglobin A1c; BMI = body mass index; SBP =Systolic blood
pressure; DBP =diastolic blood pressure; LDL-C=low density lipoprotein; HDL-C=high
density lipoprotein;GFR = glomerular filtration rate; CPR=C-reactive protein; CCT=central
cornea thickness;CP=cornea power; D=dioptr;ACD= anterior chamber depth;LT=lens
thickness;SE=spherical equivalent;DR = diabetic retinopathy.

### RT and AL

The average RT of the foveal area across the AL quartiles was 221.7 ± 25.3, 228.4 ± 34.1, 229.2 ± 29.1, and 235.9 ± 35.8 μm, respectively, and it showed an increasing trend as the AL elongated (*P* < 0.001). A decreasing trend was observed for RT as AL increased in the outer ring, the mean values for the AL quartiles being 267.8 ± 17.1, 265.8 ± 18.1, 263.3 ± 16.3, and 259.0 ± 20.7 μm, respectively (*P* < 0.001). No significant between-group differences in RT were noted in the inner ring (*P* = 0.114) and the whole macular area (*P* = 0.554) (Table 3).

**Table3.** Comparison of retinal and choroidal thickness stratified by axial length quartile among diabetic participants

| Characteristic | Axial length(mm) |  |  |  | P value |
| --- | --- | --- | --- | --- | --- |
|  | First quartile<br>(<22.84) | Second quartile<br>(22.84~23.43) | Third quartile<br>(23.43~24.06) | Fourth quartile<br>(≥24.06) |  |
| **Retinal thickness** |  |  |  |  |  |
| Central grid | 221.7±25.3 | 228.4±34.1 | 229.2±29.1 | 235.9±35.8 | <0.001 |
| Inner nasal grid | 297.8±21.5 | 301.1±21.5 | 300.7±24.3 | 302.7±27.7 | 0.069 |
| Inner superior grid | 299.5±20.3 | 302.2±21.0 | 303.0±19.1 | 302.3±22.7 | 0.142 |
| Inner temporal grid | 286.8±21.1 | 290.0±27.3 | 289.8±23.7 | 288.8±25.7 | 0.321 |
| Inner inferior grid | 295.0±20.9 | 298.3±25.0 | 299.1±22.8 | 298.6±26.7 | 0.101 |
| Average inner ring | 294.8±19.8 | 297.9±21.6 | 298.2±20.7 | 298.1±22.7 | 0.114 |
| Outer nasal grid | 284.2±18.8 | 282.8±18.6 | 280.2±19.0 | 278.8±25.6 | 0.003 |
| Outer superior grid | 269.9±18.3 | 268.0±17.5 | 265.9±19.9 | 261.2±21.9 | <0.001 |
| Outer temporal grid | 255.9±18.3 | 254.1±22.1 | 251.3±17.6 | 244.5±23.3 | <0.001 |
| Outer inferior grid | 261.3±17.7 | 258.3±20.6 | 255.8±18.3 | 251.6±25.4 | <0.001 |
| Average outer ring | 267.8±17.1 | 265.8±18.1 | 263.3±16.3 | 259.0±20.7 | <0.001 |
| Average retinal thickness | 274.7±17.6 | 275.9±19.4 | 275.0±17.6 | 273.8±21.5 | 0.554 |
| **Choroidal thickness** |  |  |  |  |  |
| Central grid | 233.6±80.2 | 215.3±81.2 | 206.6±81.9 | 169.7±81.2 | <0.001 |
| Inner nasal grid | 225.3±85.2 | 204.3±84.2 | 193.3±84.5 | 157.0±77.8 | <0.001 |
| Inner superior grid | 229.4±78.3 | 210.9±78.7 | 204.2±79.6 | 176.2±78.4 | <0.001 |
| Inner temporal grid | 219.6±69.9 | 203.8±72.3 | 198.6±73.0 | 170.7±74.5 | <0.001 |
| Inner inferior grid | 221.2±86.4 | 200.1±82.3 | 193.6±82.8 | 159.5±79.1 | <0.001 |
| Average inner ring | 223.9±76.1 | 204.8±76.0 | 197.4±76.4 | 165.8±74.5 | <0.001 |
| Outer nasal grid | 184.4±83.1 | 164.7±80.7 | 155.6±80.0 | 118.9±64.3 | <0.001 |
| Outer superior grid | 213.6±73.2 | 202.3±75.6 | 195.4±72.7 | 174.3±74.1 | <0.001 |
| Outer temporal grid | 192.7±64.1 | 178.2±63.1 | 177.7±64.0 | 157.5±65.4 | <0.001 |
| Outer inferior grid | 196.9±82.3 | 176.7±77.0 | 173.7±80.6 | 142.6±68.6 | <0.001 |
| Average outer ring | 196.9±70.0 | 180.5±68.7 | 175.6±68.5 | 148.3±62.3 | <0.001 |
| Average choroidal thickness | 213.0±72.6 | 195.1±72.3 | 188.7±72.6 | 158.5±68.8 | <0.001 |

In multiple regression models, AL was positively associated with central macular RT (β = 4.11 per mm of increase in AL, 95% CI = 2.66 to 5.56, *P* < 0.001), but negatively associated with RT in the outer ring (β = −3.37 per mm of increase in AL, 95% CI = −4.19 to −2.56, *P* < 0.001). We further analyzed associations of different AL quartiles with RT, and the results showed a positive association of the AL quartiles with RT in the macular foveal area (β = 4.90 for the second quartile, increasing to 10.3 for the fourth quartile; *P* < 0.05) and a negative association of the AL quartiles with RT in the outer ring (β = −2.75 for the second quartile, increasing to −11.5 for the fourth quartile, *P* < 0.05). No association between AL and RT in the inner ring was found in the current study (β = 0.47 per mm of increase in AL, 95% CI = −0.52 to 1.46, *P* = 0.349) (Table 4).

**Table 4.** Associations between axial length and retinal thickness among diabetic participants

|  | Central retinal thickness |  | Average retinal thickness of inner ring |  |
| --- | --- | --- | --- | --- |
|  | Model1 | Model2 | Model1 | Model2 |
| | $\beta$(95%CI) | $\beta$(95%CI) | $\beta$(95%CI) | $\beta$(95%CI) |
| **Axial length** |  |  |  |  |
| Quartiles1 | - | - | - | - |
| Quartiles2 | 5.04(0.29,9.78) | 4.90(0.20,9.60) | 1.99(-1.13,5.12) | 1.95(-1.23,5.13) |
| Quartiles3 | 4.99(0.19,9.79) | 5.54(0.79,10.3) | 1.79(-1.38,4.95) | 2.22(-0.99,5.43) |
| Quartiles4 | 11.0(6.11,15.8) | 10.3(5.48,15.1) | 0.62(-2.57,3.82) | 0.70(-2.53,3.94) |
| Per mm increased | 4.09(2.63,5.54) | 4.11(2.66,5.56) | 0.29(-0.67,1.25) | 0.47(-0.52,1.46) |

3 Modle1: Adjusted for age and sex 4 Modle2: Adjusted for age, sex, duration of diabetes, use of insulin, hemoglobin 5 A1c, BMI, SBP, hypertension,GFR, cholesterol and triglycerides.

### CT and AL

The average CT of the whole macular area was 213.0 ± 72.6 μm for the first quartile, 195.1 ± 72.3 μm for the second quartile, 188.7 ± 72.6 μm for the third quartile, and 158.5 ± 68.8 μm for the fourth quartile, indicating a decreasing trend as AL elongated (*P* < 0.001). CT tended to decrease as AL elongated in all nine subgrids (*P* < 0.05 in all cases) (Table 3). Longer eyes tended to have thinner CTs irrespective of the presence of DR (Figure 1).

**Figure1.**
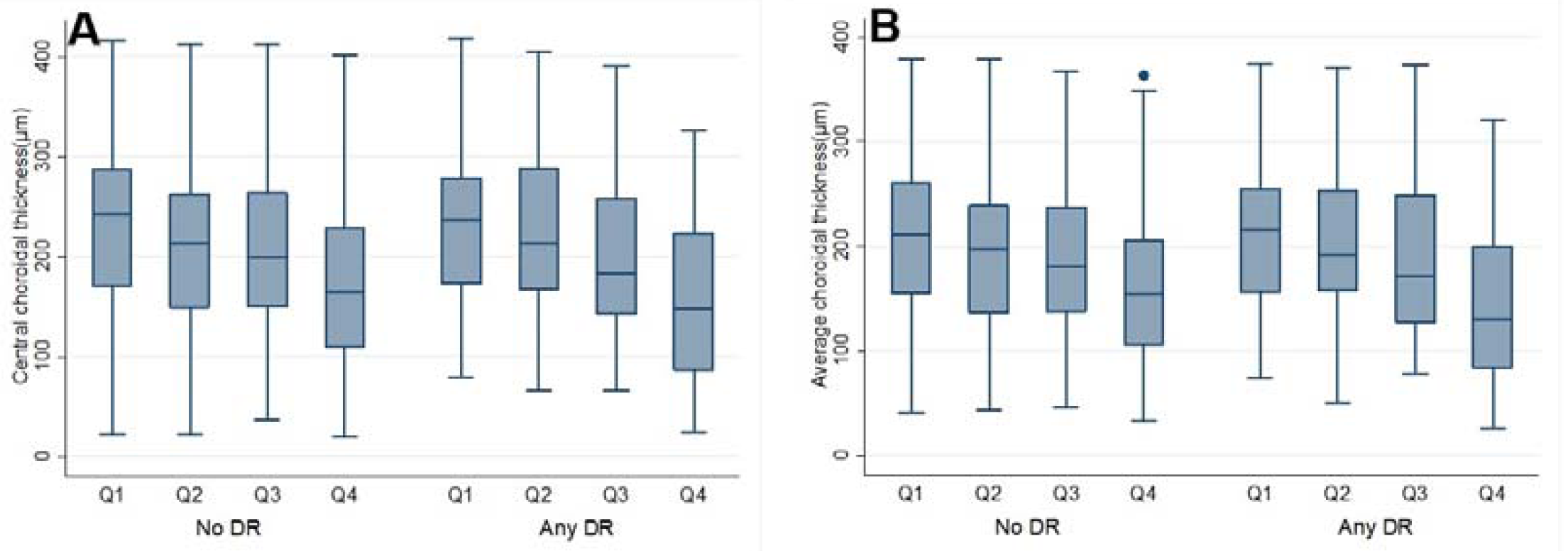
Boxplots of choroidal thickness stratified by AL quartile and DR status. **A**) central choroidal thickness **B**) average choroidal thickness. **Q1**=AL quartile1;**Q2**=AL quartile2;**Q3**=AL quartile3;**Q4**=AL quartile4.

In the multivariate regression model, a longer AL was significantly associated with a thinner CT in the foveal macular area(β = −27.4 per mm of increase in AL, 95% CI = −31.2 to −23.7, *P* < 0.001), and the impact of AL on CT increased significantly from the second quartile (β = −20.3, 95% CI = −32.2 to −8.44, *P* = 0.001) to the fourth quartile (β = −74.7, 95%CI = −86.9 to −62.5, *P* < 0.001).

The same association trends of AL and CT were detected in the outer (β = −24.6 per mm of increase in AL, 95%CI = −28.1 to −21.1, *P* < 0.001) and inner rings (β = −20.8 per mm of increase in AL, 95% CI = −23.8 to −17.7, *P* < 0.001) (Table 5).

**Table 5.** Associations between axial length and choroid thickness among diabetic participants

|  | Central choroidal thickness |  | Average choroidal thickness of inner ring |  |
| --- | --- | --- | --- | --- |
|  | Model1 | Model2 | Model1 | Model2 |
| | $\beta$(95%CI) | $\beta$(95%CI) | $\beta$(95%CI) | $\beta$(95%CI) |
| **Axial length** |  |  |  |  |
| Quartiles1 | - | - | - | - |
| Quartiles2 | -22.2(-33.9,-10.5) | -20.3(-32.2,-8.44) | -22.7(-33.5,-11.9) | -20.7(-31.7,-9.71) |
| Quartiles3 | -33.5(-45.4,-21.5) | -33.3(-45.3,-21.2) | -32.4(-43.4,-21.3) | -31.9(-43.1,-20.7) |
| Quartiles4 | -76.5(-88.6,-64.4) | -74.7(-86.9,-62.5) | -69.9(-81.1,-58.7) | -68.1(-79.4,-56.8) |
| Per mm increased | -27.9(-31.6,-24.2) | -27.4(-31.2,-23.7) | -25.1(-28.6,-21.7) | -24.6(-28.1,-21.1) |

Modle1: Adjusted for age and sex Modle2: Adjusted for age, sex, duration of diabetes, use of insulin, hemoglobin A1c, BMI, SBP, hypertension,GFR, cholesterol and triglycerides.

## Discussion

In the current study, we first documented the distribution and associations of AL with CT and RT among the participants with T2DM. The results of the same multivariate regression model indicated that AL was positively associated with the foveal RT but negatively associated with the RT of the outer ring, while the RT of the inner ring was not associated with AL. A significant negative association between AL and CT in all macular subgrids was found in the current study.

Numerous studies have demonstrated an independent relationship between RT and AL among healthy participants. Population-based studies, including the Beijing Eye Study^18^ and the Handan Eye Study^19^, found that foveal RT was positively correlated with AL. One study of 194 participants aged 6 to 17 years showed that participants with moderate to high myopia tended to have greater foveal RT than those with emmetropia or low myopia, but have thinner quadrant-specific RT in the outer ring.^20^ Another study on 80 high myopic eyes and 40 non-myopic eyes found that high myopic eyes had greater mean RT in the foveal area, but the mean RTs in the inner and outer rings of the high myopic eyes tended to be thinner than those in the control eyes.^21^ Research on 336 healthy young Korean subjects aged 19 to 25 years showed that the foveal RT increased while the RT of the inner/outer ring decreased as the degree of myopia/AL increased.^22^ The Sydney Childhood Eye Study reported that the RT of the outer ring, and not that of the foveal area, thinned as AL increased among 1,534 children aged 6 years.^23^ However, some other studies indicated no association between AL and foveal RT.^24^ Research on RT among diabetic patients is limited. Our study is the first to show that the foveal RT increases with ocular elongation, while the RT of the outer ring thins with longer AL among Chinese diabetic patients.

Using Spectralis HRA+OCT, the peripheral RT outside the ETDRS grid was also verified to be linearly associated with AL among 50 healthy participants.^25^ The Beijing Eye Study showed that the mean thickness of the inner and outer nuclear layers at the equator and the midpoint of the equator decreased significantly with AL elongation in enucleated eyes.^18^ The findings of the current study and previous works show that RT thickened in the foveal area and thinned in the outer ring with AL elongation, mirroring the theory that myopia-related elongation of the globe walls primarily occurs in the midperiphery or the equatorial region.^26^

The choroid provides oxygen and nourishment to the outer retinal layers, thus regulating temperature, and it alone provides blood supply to the macular avascular fovea.^27^ Many studies have suggested the key role of CT in DR pathologies.^11, 12, 28^ Thus, it is crucial to confirm the impact of AL on CT for diabetic eyes. Our finding that AL elongation is accompanied with thinning of the foveal CT among diabetic patients is consistent with the results of the Beijing Eye Study,^11^ which included 269 diabetic participants. The Kailuan Diabetic Retinopathy Study indicated that subfoveal CT decreased with AL elongation among 1,154 diabetic participants.^5^ However, both these studies did not analyze the relationship between AL and CT in other macular subgrids. The reverse relationship between AL and foveal CT was also confirmed in high myopic eyes,^29^ young health adults,^30^ those with angle closure,^31^ and population-based studies.^32^ Attenuation of CT is associated with decreased choroid–retinal blood flow and reduced metabolic demand, which may impart a protective mechanism to myopic eyes, preventing them from developing DR. Thus, further studies evaluating the associations between AL and choroid blood flow are warranted. Additionally, our work indicated that AL has an unbalanced impact on CT in different subregions; the impact of AL on CT was most significant in the foveal area (β = −27.4), and this impact was attenuated in the outer ring (β = −20.8). A previous study on 150 young health adults also indicated a slower decrease rate of CT with AL elongation in the outer subfields.^30^ Further investigations are thus needed to confirm if these region-wide disparities concerning AL and CT have any impact on the presence and progression of DR.

The current study enjoyed several advantages over previous similar works. It used a large study sample with a standard study protocol, as well as strict exclusion criteria (e.g., intraocular surgeries or other retinal diseases), which prevented confounder factors that could have affected the reliability of the results. The relatively higher resolution scanning depth and speed of the SS-OCT made the measurements more reliable. However, the study also suffered from several limitations. First, the participants for the study were recruited from communities in Guangzhou only. Thus, the conclusions cannot be generalized to other areas. Second, unlike longitudinal studies, the current cross-sectional study cannot provide answers regarding cause and effect on the association between AL and RT, and AL and CT. Third, the participants in our study underwent OCT scanning at any time of the day, which may have generated a quantitative bias; a previous study indicated a diurnal variation of approximately 20 to 30 μm for CT when using OCT measurements.^33^

In conclusion, ocular elongation is accompanied by retinal thinning of the outer ring and retinal thickening of the foveal area. CT tended to thin with AL elongation among diabetic participants. Further longitudinal studies are needed to confirm this relationship between AL and RT, and AL and CT.

## Data Availability

The availability of all data referred to in the manuscript and note links below.

## Acknowledgment

None

## Footnote

The authors have completed the STROBE reporting checklist

## Reference

1. Yau JW, Rogers SL, Kawasaki R, et al. Global prevalence and major risk factors of diabetic retinopathy. Diabetes care 2012; 35(3): 556–64.

2. Zheng Y, He M, Congdon N. The worldwide epidemic of diabetic retinopathy. Indian journal of ophthalmology 2012; 60(5): 428–31.

3. Man RE, Sasongko MB, Sanmugasundram S, et al. Longer axial length is protective of diabetic retinopathy and macular edema. Ophthalmology 2012; 119(9): 1754–9.

4. Lim LS, Lamoureux E, Saw SM, Tay WT, Mitchell P, Wong TY. Are myopic eyes less likely to have diabetic retinopathy? Ophthalmology 2010; 117(3): 524–30.

5. Wang Q, Wang YX, Wu SL, et al. Ocular Axial Length and Diabetic Retinopathy: The Kailuan Eye Study. Investigative ophthalmology & visual science 2019; 60(10): 3689–95.

6. He J, Xu X, Zhu J, et al. Lens Power, Axial Length-to-Corneal Radius Ratio, and Association with Diabetic Retinopathy in the Adult Population with Type 2 Diabetes. Ophthalmology 2017; 124(3): 326–35.

7. Quigley M, Cohen S. A new pressure attenuation index to evaluate retinal circulation. A link to protective factors in diabetic retinopathy. Archives of ophthalmology 1999; 117(1): 84–9.

8. Stefansson E. Ocular oxygenation and the treatment of diabetic retinopathy. Survey of ophthalmology 2006; 51(4): 364–80.

9. Querques G, Lattanzio R, Querques L, et al. Enhanced depth imaging optical coherence tomography in type 2 diabetes. Investigative ophthalmology & visual science 2012; 53(10): 6017-24.

10. Regatieri CV, Branchini L, Carmody J, Fujimoto JG, Duker JS. Choroidal thickness in patients with diabetic retinopathy analyzed by spectral-domain optical coherence tomography. Retina 2012; 32(3): 563–8.

11. Xu J, Xu L, Du KF, et al. Subfoveal choroidal thickness in diabetes and diabetic retinopathy. Ophthalmology 2013; 120(10): 2023–8.

12. Lains I, Talcott KE, Santos AR, et al. Choroidal Thickness in Diabetic Retinopathy Assessed with Swept-Source Optical Coherence Tomography. Retina 2018; 38(1): 173–82.

13. Michalewski J, Michalewska Z, Nawrocka Z, Bednarski M, Nawrocki J. Correlation of choroidal thickness and volume measurements with axial length and age using swept source optical coherence tomography and optical low-coherence reflectometry. BioMed research international 2014; 2014: 639160.

14. Hirata M, Tsujikawa A, Matsumoto A, et al. Macular choroidal thickness and volume in normal subjects measured by swept-source optical coherence tomography. Investigative ophthalmology & visual science 2011; 52(8): 4971–8.

15. Alonso-Caneiro D, Read SA, Collins MJ. Automatic segmentation of choroidal thickness in optical coherence tomography. Biomedical optics express 2013; 4(12): 2795–812.

16. Revised Grading Definitions for the NHS Diabetic Eye Screening Programme. Available at: https://www.gov.uk/government/publications/diabetic-eye-screening-retinal-imagegrading-criteria. Accessed June 19, 2015.

17. Olgun ME, Altuntas SC, Sert M, Tetiker T. Anemia in Patients with Diabetic Foot Ulcer: Effects on Diabetic Microvascular Complications and Related Conditions. Endocrine, metabolic & immune disorders drug targets 2019; 19(7): 985–90.

18. Jonas JB, Xu L, Wei WB, et al. Retinal Thickness and Axial Length. Investigative ophthalmology & visual science 2016; 57(4): 1791–7.

19. Duan XR, Liang YB, Friedman DS, et al. Normal macular thickness measurements using optical coherence tomography in healthy eyes of adult Chinese persons: the Handan Eye Study. Ophthalmology 2010; 117(8): 1585–94.

20. Chen S, Wang B, Dong N, Ren X, Zhang T, Xiao L. Macular measurements using spectral-domain optical coherence tomography in Chinese myopic children. Investigative ophthalmology & visual science 2014; 55(11): 7410–6.

21. Wu PC, Chen YJ, Chen CH, et al. Assessment of macular retinal thickness and volume in normal eyes and highly myopic eyes with third-generation optical coherence tomography. Eye 2008; 22(4): 551–5.

22. Hwang YH, Kim YY. Macular thickness and volume of myopic eyes measured using spectral-domain optical coherence tomography. Clinical & experimental optometry 2012; 95(5): 492–8.

23. Huynh SC, Wang XY, Rochtchina E, Mitchell P. Distribution of macular thickness by optical coherence tomography: findings from a population-based study of 6-year-old children. Investigative ophthalmology & visual science 2006; 47(6): 2351–7.

24. Wang J, Gao X, Huang W, et al. Swept-source optical coherence tomography imaging of macular retinal and choroidal structures in healthy eyes. BMC ophthalmology 2015; 15: 122.

25. Wenner Y, Wismann S, Preising MN, Jager M, Pons-Kuhnemann J, Lorenz B. Normative values of peripheral retinal thickness measured with Spectralis OCT in healthy young adults. Graefe’s archive for clinical and experimental ophthalmology = Albrecht von Graefes Archiv fur klinische und experimentelle Ophthalmologie 2014; 252(8): 1195–205.

26. Vurgese S, Panda-Jonas S, Jonas JB. Scleral thickness in human eyes. PloS one 2012; 7(1): e29692.

27. Nickla DL, Wallman J. The multifunctional choroid. Progress in retinal and eye research 2010; 29(2): 144–68.

28. Kim JT, Lee DH, Joe SG, Kim JG, Yoon YH. Changes in choroidal thickness in relation to the severity of retinopathy and macular edema in type 2 diabetic patients. Investigative ophthalmology & visual science 2013; 54(5): 3378–84.

29. Flores-Moreno I, Lugo F, Duker JS, Ruiz-Moreno JM. The relationship between axial length and choroidal thickness in eyes with high myopia. American journal of ophthalmology 2013; 155(2): 314–9 e1.

30. Tan CS, Cheong KX. Macular choroidal thicknesses in healthy adults--relationship with ocular and demographic factors. Investigative ophthalmology & visual science 2014; 55(10): 6452–8.

31. Huang W, Wang W, Gao X, et al. Choroidal thickness in the subtypes of angle closure: an EDI-OCT study. Investigative ophthalmology & visual science 2013; 54(13): 7849–53.

32. Wei WB, Xu L, Jonas JB, et al. Subfoveal choroidal thickness: the Beijing Eye Study. Ophthalmology 2013; 120(1): 175–80.

33. Tan CS, Ouyang Y, Ruiz H, Sadda SR. Diurnal variation of *choroidal thickness in normal*, healthy subjects measured by spectral domain optical coherence tomography. Investigative ophthalmology & visual science 2012; 53(1): 261–6.

